# Deficiencies in accessibility to point-of-care (POC) diagnostics in PMTCT services in rural primary health care clinics in Zambia: Implementation Science perspective

**DOI:** 10.1101/2024.07.11.24310263

**Authors:** J Katoba

**Affiliations:** Discipline of Public Health Medicine, School of Nursing and Public Health, University of KwaZulu-Natal, South Africa

## Abstract

Point-of-care (POC) innovations are healthcare interventions that have great potential to improve diagnostic capacities of healthcare systems in low- and middle-income countries (LMICs). From the implementation science perspective, this study explores perceptions of key stakeholders on deficiencies in accessibility of POC diagnostics in the context of prevention of mother-to-child transmission (PMTCT) services in Zambia.

**Methods:** A retrospective qualitative evaluation was conducted to understand factors that influence POC diagnostic implementation in rural primary healthcare (PHC) settings. The study was conducted among key implementing stakeholders in Zambia. In-depth interviews were held with 11 purposefully selected key informants, including clinical officers, midwives, nurses, environmental health technicians (EHTs), government and private health officials. The interviews were audio-recorded and transcribed verbatim. Following coding, thematic content analysis was applied and the main emerging themes were analysed through the lens of the Consolidated Framework for Implementation Research (CFIR). The CFIR was identified as the most appropriate model to interpret our findings.

**Results:** Factors influencing implementation were represented in all five domains. Major constructs as facilitators were the relative advantage of the intervention, external partnership, education and training, knowledge and belief, self-efficacy, and engagement of champions. Barriers were mainly found in the outer and inner settings, including constraints in financial resources, supply chain challenges resulting in stock-outs, insufficient human resources leading to increased workload, and other infrastructural issues like space limitations and lack of electricity in most rural primary healthcare settings.

**Conclusions:** The study identified key determinants that supported or hindered the implementation of POC diagnostics in the rural PHCs. Greater efforts are needed to overcome barriers at multi-sectoral level for effective implementation while leveraging on facilitators through a health system strengthening approach. These findings are key to informing future implementations, sustainability and scale-up of POC diagnostics interventions.

## Background

Preventing childhood and maternal diseases, including HIV, is an important strategy that can reduce pediatric HIV, improve child survival and a mother’s health [1]. However, in LMIC, there is a lack of testing technologies in primary healthcare systems for many diseases other than diagnosing HIV in adults [2]. This can lead to missed opportunities for delivery of health services and low opportunities for paediatric case identification and provision of paediatric antiretroviral therapy (ART).

Although enormous progress has been made so far in prevention of mother-to-child transmission (PMTCT) programmes, approximately 90% of new HIV cases in children < 15 years old still get infected through mother-to-child (MTCT) annually, due to undetected new infections during pregnancy/breast feeding [3]. One of the global health approaches to build upon the gains of PMTCT and improve maternal and child health is improving accessibility to point-of-care (POC) diagnostic services during antenatal care (ANC) and perinatal care [3].

POC diagnostics are innovations with potential for early diagnosis and timely linkage to care for patients. In addition to these possible gains, POC technologies provide a window of opportunity for providing integrated healthcare services in PMTCT/MNCH [2]. As described elsewhere, the goal of POCT is to provide a quick test result for immediate clinical decisions to improve patients’ health outcomes [4]. Other benefits of integrated POCT services have also been reported in previous publications, including cost-effectiveness, increased testing uptake and treatment rates [4]. The Zambian prevention of mother-to-child transmission guidelines, which are aligned with the World Health Organization’s (WHO) recommendation on provision of integrated services during routine antenatal care, require testing for infections including HIV, syphilis, malaria, TB and other ailments as part of their efforts to contribute to improved maternal and neonatal outcomes in Zambia [5]. Despite all these strategies, screening and treatment of infections is still not universal; thus, onward transmission of infections from mother-to-child still occur [6, 7].

WHO has made recommendations for POCT and has prequalified *In Vitro* diagnostic products to ease the service delivery gap in areas with unmet diagnostic capabilities [8]. Hence, use of POC tests in settings burdened with malaria, HIV, syphilis, TB and lower respiratory tract infections are anticipated to save more than one million lives yearly [9]. Furthermore, WHO has published a list of essential *in-vitro* diagnostics needed for diagnosing most common illnesses, as well as a number of global priority diseases, including HIV [10, 11].The list provides guidance and reference for the development of country-specific lists of essential diagnostics. Thus, these simple POC rapid diagnostics on regional and national programmes hold immense promise for tackling various public health programmes. However, access to these innovations across all settings is required to realize their benefits. For instance, access to easy-to-use diagnostics for antenatal syphilis is expected to save at least 138 000 lives and avert more than 148 000 stillbirths annually [9, 12].

Diagnostic services are a critical step in the PMTCT cascade from diagnosis to ART initiation and viral suppressions [13]. Subsequently, the ”test and treat” approach [14], a care package of POC diagnostic interventions in the context of PMTCT and other specific tests, has been implemented and offered in public healthcare settings of Zambia by the Ministry of Health and its cooperation partners [5]. However, despite these approaches, there is still limited information on factors contributing to the implementation process, or information elaborating on the context in which these interventions are designed or conducted in rural primary care settings in Zambia. We adopted an implementation research perspective to systematically gain insights into factors contributing to the success of POC diagnostic intervention programme implementation. Implementation research intends to understand why, in what context, and for who interventions work in “real-world settings”[15]. The aim was to explore reasons for deficiencies in accessibility of POC diagnostics in PMTCT services in Zambia. The findings may support efforts and build on knowledge of what works, and of the underlying contextual factors for effective implementation of POC diagnostics in rural primary healthcare practices in Zambia and the region.

## Methods

### Study setting and context

The study reported here was nested within a larger study entitled “Accessibility of point-of-care diagnostics for prevention of vertical disease transmission in rural primary healthcare clinics in Zambia”. This study was conducted in different settings, including Central Province of Zambia. Central Province is one of the 10 provinces of Zambia described predominantly as rural. Rural areas as defined elsewhere are communities that are separated from central clusters (i.e., towns) and may be deprived of modern amenities such as general supply of electricity and/or piped water and ease of access to medical care, schools and recreation facilities [16]. Some surface areas in the province have poor terrain and are covered by rivers, wetlands, and hills. For example, there is a major wetland known as Lukanga swamp, which is a permanently swampy area in the province covering an area of approximately 2 600 km^2^ in total. In Zambia, government-owned public health facilities are the main providers of health services in urban and rural areas. Public healthcare delivery is organised generally in a referral system comprising: 1^st^ level (health posts, health centres and district hospitals); 2^nd^ level (provincial and general hospitals); and 3^rd^ level at tertiary level (teaching and central Hospitals). Primary healthcare and preventive health services are provided at rural health posts, rural health centres and district hospitals. The demand for these services is highest from rural populations. Rural health posts service about 3 500 people within a 5 km radius, followed by rural health centres that cater for 10 000 people located within a radius of 30 km [17]. The provincial map is captured in the national map of Zambia in Figure 1.

**Figure 1:**
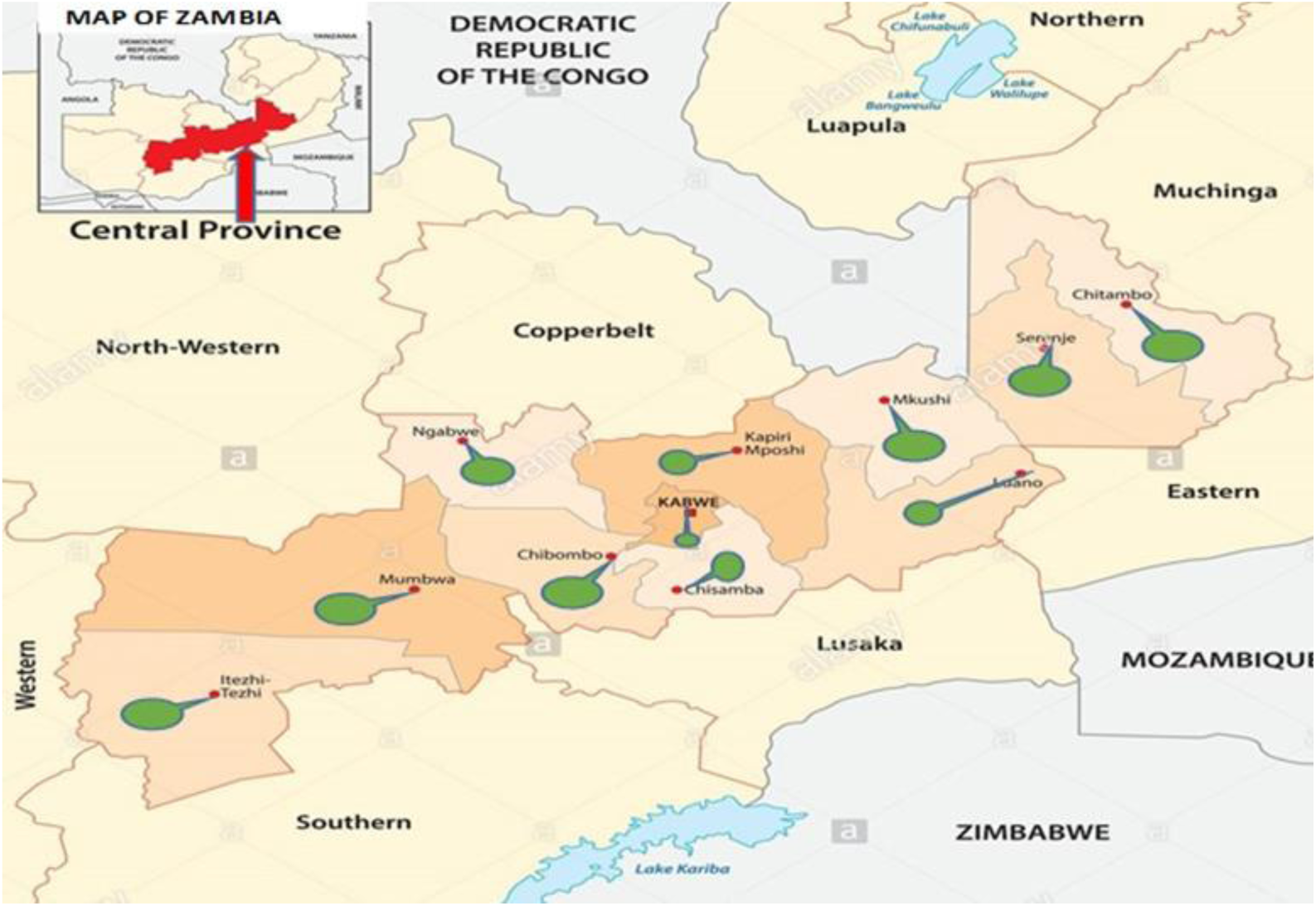
National map of Zambia showing Central Province with 11 districts highlighted in green

### Study design

A retrospective evaluation with a qualitative design was conducted to understand the implementation context of POC diagnostic services in rural primary health care facilities (PHCs). One of the specific research objectives was to understand facilitators and barriers to POC diagnostics implementation.

### Study participant recruitment

The study used purposeful sampling of several key stakeholders (i.e., HCWs/providers at PHCs, health officers from government and the private sector) involved in the implementation of POC interventions across different sectors of healthcare delivery. Since the majority of HCWs are POC end-users, they have sufficient experience to comment on the challenges of access to POC testing at their facilities. The government and private sector officials provide additional insight at a higher level because of their involvement in health care implementation programmes. HCWs were identified and recruited to participate in the study through a baseline survey conducted to determine accessibility of POC among HCWs stationed in rural PHCs. The HCWs were from PHCs that showed low accessibility of POC diagnostics. Clinics with low accessibility were defined as those below the overall average level of availability, based on the WHO list of essential tests per facility. In each of the selected health facilities, facility-in-charges were purposively selected for in-depth interviews. These HCWs included nurses, midwives, and clinical officers. Key persons from the district health sector and healthcare workers at facility level assisted in identifying other stakeholders who were involved in POC diagnostics. These stakeholders (government health officials and private health sector) were purposively selected and included as study participants. A total of 11 key informants were invited and consented for recruitment into the study. These included eight HCWs from primary healthcare facilities (clinical officers, nurses and community health assistants) and three stakeholders (two government health officials at national and district level and one official from the private health sector). All those who were approached attended the interviews. The composition of the interviews and their characteristics are summarised in Table 1.

**Table 1.**
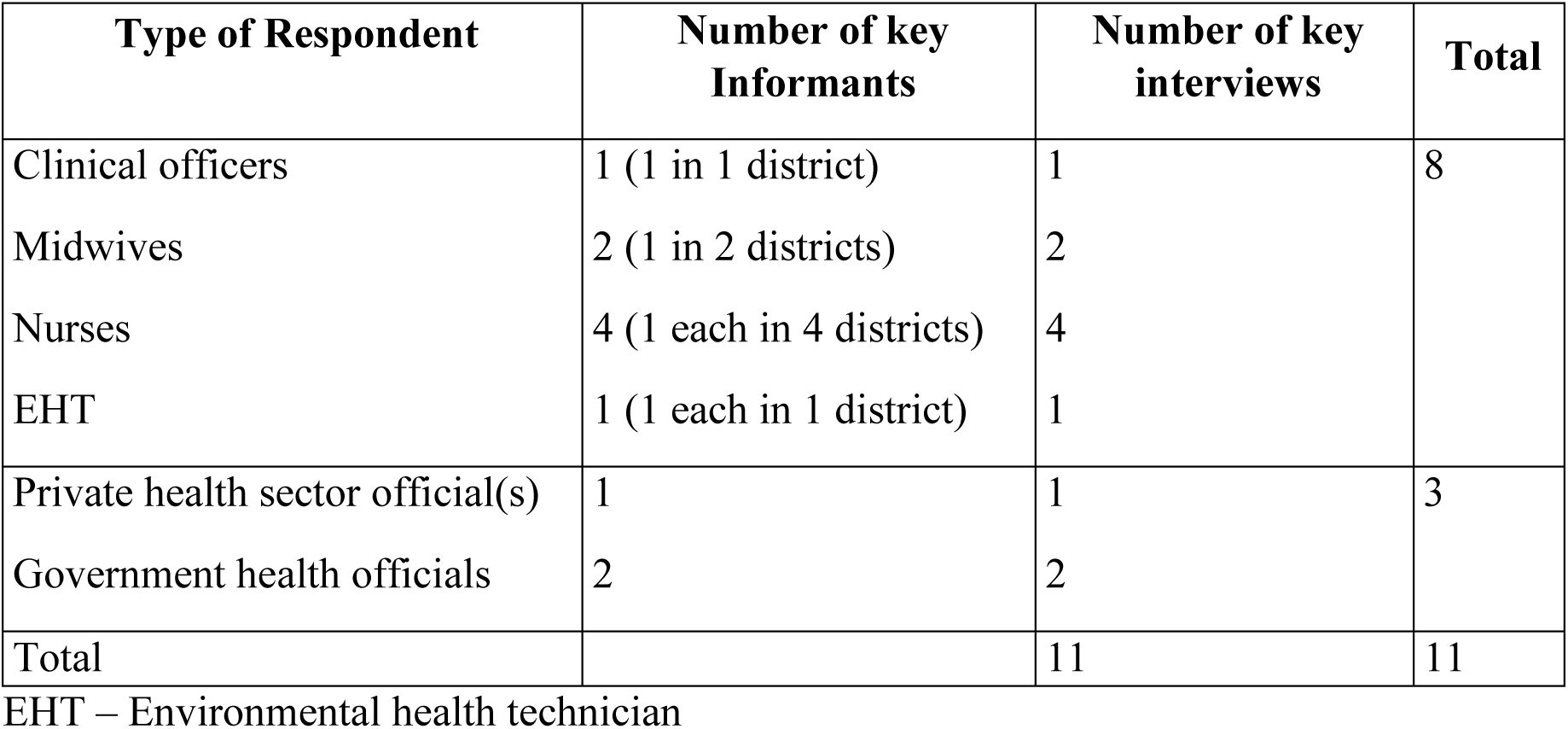
Composition of in-depth interviews and characteristics of the key informants.

| Type of Respondent | Number of key Informants | Number of key interviews | Total |
| --- | --- | --- | --- |
| Clinical officers | 1 (1 in 1 district) | 1 | 8 |
| Midwives | 2 (1 in 2 districts) | 2 |  |
| Nurses | 4 (1 each in 4 districts) | 4 |  |
| EHT | 1 (1 each in 1 district) | 1 |  |
| Private health sector official(s) | 1 | 1 | 3 |
| Government health officials | 2 | 2 |  |
| Total |  | 11 | 11 |

EHT – Environmental health technician

**Table 2.** Contextual factors affecting implementation of POC diagnostics by CFIR Domain. [19].

| **DETERMINANTS** |  |  |
| --- | --- | --- |
| **CFIR DOMAIN/CONSTRUCT** | **Facilitators** | **Barriers** |
| **INTERVENTION CHARACTERISTICS** | - Ease of use<br>- Usefulness<br>- No requirement for laboratory Infrastructure<br>- Rapid Turnaround time results<br>- Reduced referral and patient's travel costs<br>- Improved patient care and management<br>- Alignment with national and international policy on public health problems |  |
| **OUTER SETTING** | External partnerships with NGOs | - Financial resource constraints<br>- Supply chain challenges: (Complicated procurement systems, delays, poor inventory, quantification processes, logistics and insufficient storage) |
| **INNER SETTING** | - Sufficient knowledge on rapid testing<br>Acceptability (High willingness to accept diagnostic technologies)<br>- Good initial staff training<br>- Desire for on-going training<br>- Involvement of leadership of all implementers | Inadequate resources and Infrastructure<br>- Staffing dynamics (staff shortages)<br>- Supply stock-outs<br>- Extra workload<br>- Power supply challenges<br>- Transport challenges<br>- Space restrictions<br>- Technological resource (computers and internet) limitation |
| **INDIVIDUAL CHARACTERISTICS** | - Acquaintance with knowledge and skills to offer and screen using rapid tests<br>- Initial training<br>- Confidence and competency<br>- Positive attitudes towards POC testing<br>- Motivation and enthusiasm of providers | - Inconsistency training |
| **IMPLEMENTATION PROCESS** | - Involvement of relevant stakeholders such as nurses (champions) for their role as users | - Low level decision making on POC selection |

### Data collection process

Data was collected using a semi-structured interview guide, adapted from similar published studies [18]. The interview guide was initially pilot-tested in similar facilities not included in the study, and adjustments were made based on from pilot respondents. In-depth interviews with stakeholders who were key informants were conducted through phone call or face-to-face in English, because the participants were proficient and comfortable in the language. The face-to-face interviews were delivered in health facilities and locations where other informants involved in POC diagnostics implementation served. Key informants involved in implementation were interviewed in order to elicit information regarding barriers and enablers to POC diagnostics services in rural PHCs in Zambia, particularly in Central Province. In this study, stakeholders are defined as healthcare providers involved in patient testing and management, such as doctors, nurses, clinical officers, community health assistants, laboratory staff in rural PHCs and other health officials from public and private health sector involved in the implementation/delivery of diagnostics at various levels of healthcare delivery in the country. The interviews lasted approximately 30 minutes and were tape recorded. Interviews were conducted until saturation was reached. Further interviews were discontinued when saturation was reached (i.e., when no new information emerged).

### Data management and analysis

The interviews were recorded and afterwards transcribed verbatim. The transcripts were checked thoroughly several times to guarantee accuracy and reliability of the interviews. To become familiarised with the data, transcripts were read and re-read line by line. Due to a relatively small number of interviews, inductive analysis was done manually. Codes were identified *a priori* mirroring constructs from the Consolidated Framework for Implementation Research (CFIR) [19]. After a few transcripts were analysed, no new codes emerged, an indication that there was data saturation and therefore there was no need for further data collection. Major and additional sub-themes that emerged from the data formulated the final themes. Following thematic content analysis, based on the final themes, CFIR [19] was applied to fully understand the implementation dynamics and was chosen as the most suitable model to provide theoretical basis to interpret our findings. The CFIR is a comprehensive framework designed to systematically identify multilevel context factors that are likely to influence implementation and effectiveness of a given intervention [20, 21]. This framework consists of 39 specific constructs organised across five domains, all of which interact to influence implementation. The following are the five major domains:

- *Intervention characteristics (e.g., toolkit relative advantages, adaptability etc.);*
- *Outer setting (e.g.,. resources, external healthcare system policies and incentives);*
- *Inner setting (e.g., structural and facility characteristics);*
- *Characteristics of individuals involved (knowledge, self-efficacy); and*
- *Implementation process (e.g., planning and engaging stakeholders, dissemination)* [20, 22].

The CFIR was selected for this study because of its flexibility across a wide range of applications at any stage of implementation (pre, during, post) [23].

### Ethical approval

The study was approved by the University of KwaZulu-Natal, Biomedical Research Ethics Committee (BREC Ref No: BE650/17) and the University of Zambia, Biomedical Research Ethics Committee (Approval Number: 012-02-18). Further permission and authority to collect data from the health facilities was obtained from the Zambia National Health Research Authority (ZNHA) and Provincial and District Health Office. Written informed consent was obtained from all participants prior to the study. No participant withdrew their consent. Participation in the study was voluntary, and all participants were free to withdraw at any time. All participants were assured of confidentiality and anonymity during the interview, transcription, and analysis stages.

## Results

### Participants’ characteristics

The characteristics of respondents are presented in Table 1. A total of 11 key informants participated in the study. Eight were facility-based health providers /POC-end users (clinical officers, midwives, nurses and environmental health technicians) and three public and private health officials. Nurses represented 50% of the HCWs interviewed in PHCs. All HCWs interviewed had work experience of more than one year prior to the interviews.

### Contextual factors that influenced implementation of the intervention

The findings are presented in the context of CFIR domains embedding the constructs that emerged [19]. We present key themes that informants felt either supported or hindered implementation of a care package of POC (HIV, syphilis, malaria) interventions. Below, key themes are presented as facilitators and barriers, and summarised in Table 1. Selected direct quotes from the interviews to illustrate the findings are provided and cited throughout the results.

### Intervention characteristics

Relative advantages of POC diagnostics were common and strong themes that emerged across all stakeholders. The perceived advantages included rapid testing, improved turnaround time, prompt detection of infections to improve patient care and management, reduced travel costs for patients, and reduced referrals. Facility-based health informants reported increased confidence in POC testing and felt empowered to provide better care without having to refer all patients for diagnosis. These aspects served as facilitators to the implementation of the intervention due to non-complexity of the intervention.

> *“The advantage for point-of care testing is that I think they lessen the burden on the patients, especially when it comes to travel costs, because patients don’t have to travel somewhere else, where there are labs for them to access that service, so when they come to this facility, we can be able to provide the tests they need” (Participant 4)*

> *“Because first of all, it quickens the process of diagnosis and proper management. Because the quicker you diagnose the quicker you manage and the more you prevent diseases escalating to the higher levels. If I manage a case of malaria at a health centre or health post, it means that it will not go to the hospital and if it doesn’t go to the hospital, they are not going to use a cannula, they are not going to use a needle, they are not going to use an injectable drug and these are all these are expensive collectively if you put them together, so that’s why a point of care test should be available at any health centre especially rural areas. The goal is to prevent referrals and reduce on health care expenditure in the hospitals” (Participant 9).*

### Outer settings

The constructs which were assessed under this domain were resources, networking and external policies.

Under this domain “cosmopolitanism” was the main construct related to networking and was reported as a facilitator to implementation. Cosmopolitan describes the degree to which the implementing team is externally networked, with connections to other entities assumed to promote an implementation [19]. External support from non-government affiliated partners such as John Snow, Inc (JSI) served as a catalyst to advance the national provision of test kits, particularly for HIV services. Involvement of donors and other UN agencies such as USAID and the U.S. President’s Emergency Plan for AIDS Relief (PEPFAR) also added to multi-sectoral coordination efforts and facilitated programme implementation.

> *“We are partners with Ministry of Health, like I said we just do our part which is, mainly assisting Ministry of Health do the distribution and where there are gaps we come in and assist”…”There are so many of us, like USAID and PEPFAR that provide test kits especially relating to HIV and AIDS malaria. We provide technical support and facilitate delivery of those commodities just to make sure that the commodity availability and we make sure we do self-delivery. Other test kits are also provided by USAID and other partners”. (Participant 7)*

Under this outer domain, constructs identified to hinder implementation success were resources and external policy related to supply chain challenges.

#### Financial resources constraint

Availability of resources such as funding was a construct highlighted as critical in determining the level of POC diagnostics availability. Due to limited financial resources to support health care, the ability to deliver health interventions is restricted, particularly in rural settings. Key informants noted that POC diagnostic tests needed to be available at every service delivery point in order to provide quality health care. However, other informants mentioned that certain tests needed to be prioritised due to challenges in resources. For example, HIV rapid tests as compared to CD4 POC count was strongly cited as one of the tests that needed to be prioritised so that many patients are initiated on ART to reduce transmission.

> *“So, the major reason if we have to sum up why we may not have everything at the facility is a “resource envelope”, the resource envelope if not managed at the moment, is the more reason why we have thought to have the “social health insurance” (participant 9) “I said where you have a lot of competing priorities sometimes it becomes very difficult to prioritise. So maybe that’s why the number of facilities they run out of the tests. I think if we had a lot of resources at our disposal, we would make sure that the facilities are well stocked, and we have all the tests needed”. (Participant 5)*

> *“It depends on the resources; we would want to have everything that we have listed down at the point of care but there are times we run out of resources and so have to run around to look for alternatives”. (Participant 5)*

#### Supply chain challenges

The challenges associated with the supply chain system were highlighted as obstructing implementation. Issues relating to stock-outs were frequently reported as a key hindrance to access to POC diagnostic services in PHCs. It was further acknowledged that challenges in aspects such as forecasting, complicated and inefficient procurement system, inventory control, inadequate storage facilities, distribution and logistics management were a hindrance to implementation. Furthermore, issues of the supply not always meeting demand, lead time and unrealistic timeframe are some of the procurement challenges that were reported as causing substantial delays in supplies. Other highlighted challenges related to delays in supply and distribution included delays in inventory reports, and logistical challenges coupled with vehicle breakdowns in some areas with poor terrain.

> *“Lead time is the time that we expect the commodities to be made available in use at a facility or service delivery point, from the time they are ordered. So, if the lead time is longer, it means that facility will eventually run out of supplies”. (Participant 7)*

### Inner setting

The construct assessed under this domain was readiness for implementation. This included leadership engagement, access to available resources and access to knowledge and information as factors that influenced implementation.

### Supportive leadership

Another critical element that facilitated implementation was active leadership. The support through leadership at national and district level and facility in-charges actively participating in the implementation and use of POC devices was found to be valuable to implementation through policy, guidelines and material resources. This supportive role was cited as important at individual and institutional level. Government support also facilitated meaningful engagement with relevant stakeholders, including champions and NGOs.

> *“I think it’s important to bring everyone on board. There are a lot of us who don’t have a clue about different types POC testing or use it because they were not brought in in the beginning. Then if you are engaged and involved and understand the benefits, I think it makes a difference”. (Participant 1)*

#### Available resources

In terms of available resources, stock-outs, functional equipment, human resources, space restrictions, power supply and logistical resources were barriers identified under this domain.

##### Insufficient stock

Generally, facility-based informants reported shortages and frequent stock-outs as a major challenge to regular provision of POC diagnostic services. Although they were not able to exactly state why shortages occurred, they indicated that most facilities received support to ensure availability of HIV and malaria RDTs. However, the level of support for other tests that were also needed was not uniform. These included simple POC tests for diagnosing HIV in infants in rural facilities. All facilities were currently using Dried Blood Spots (DBS) testing, which was reportedly associated with sample transportation and delayed results challenges. Occasional stock-outs of syphilis tests of more than one year were reported in one facility. Other informants reported expired products, delays in supplies and sometimes inadequate supplies of commodities.

> *“We don’t have a problem with Malaria RDT. But for syphilis, supplies have been there but not always. For HIV, we usually have, though at times we run out” (Participant 3)*

> *“I think stock-outs are just supply-chain or procurement issues yes, because even if you do order, it takes a long time to receive them from the district and if you go to the district to order, you find it’s not there”. (Participant 3)*

> *“Another barrier is … In terms of TB and POC for testing infants, we have no access, we have to refer our clients and we get to know the results of a patient a little bit too late. So, knowing the results after some time is not good, especially for the positive clients and babies”. (Participant 6)*

Lack of or faulty equipment/components for glucose and haemoglobin tests, for example, were reported as a challenge. Participants mentioned that even if requests were made for components such as micro-cuvettes and strips needed to measure glucose and haemoglobin respectively, they were mostly out of stock. Providers further expressed disappointment and frustrations because they were not also supplied with a wide range of POC tests such as Hepatitis and TB Gene X-pert.

> *“Because we might order the diagnostic tests, they tell us they are out of stock, maybe the district also doesn’t have, medical stores also don’t have, and so medical stores should have these tests every day. Actually, they should be stocking these tests so that they bring them over”. (Participant 1)*

##### Inadequate staffing

Insufficient health personnel was cited as a major barrier to implementation. Access to adequate human resources was critical to successful implementation. Due to shortage of staff or limited trained personnel, increased workload was a challenge at facilities. Insufficient workforce, particularly for inventory, was mentioned as a challenge and this posed a barrier to introducing more additional tests. An increased number of additional tests with fewer providers would not only increase workload, but also waiting time for patients. The need for additional staff was reported because they felt that they were already understaffed and were overloaded with work.

##### Space limitations

Facility-based informants expressed concerns about inadequate space as a barrier. They noted that there wasn’t enough room where more tests could be done if they were supplied. Other factors including issues of privacy and confidentiality were identified as a source of concern due to limited space.

> *“The clinic needs to be expanded so that we have room, where those tests if at all we are supplied can be taking place from. Otherwise, the room is not enough, and this is a challenge. We need at least two or three rooms where we can be operating from”. (Participant 6)*

> *“Even the space itself as you can see is small. This is a screening room that is the MCH room there, the other room is labour ward, we have got only these three rooms, so it’s not enough, plus we need more staff, we are under-staffed and sometimes the work is too much, so this is another challenge”. (Participant 8)*

> *“Just looking at the state of our facility if it can be extended so that more services and tests can be provided. But like this, it’s only three rooms, so sometimes privacy and other things, is a challenge also.” (Participant 2)*

##### Transportation/logistical challenges

Informants at clinics shared that logistics were a challenge because most facilities lacked official vehicles or fuel, and this was further complicated with poor terrain in some places. Regular supply and provision of reagents and test kits required availability of transport at PHC facilities. This forced facility staff to use their own means of transport to respective district health offices to collect supplies. They mentioned that they needed vehicles or motor bikes to support daily facility activities. Facilities that were provided with motor bikes, however, noted that despite the long travel distance to the district health office, the situation had changed because it was easy for them to collect supplies whenever they run out. Having motor bikes was viewed as a catalyst for promoting intervention implementation.

> *“I will say the challenge is transport to go to the district office to go and collect those reagents. You find that, you need those things, but you don’t have transport to go and collect them and this is a barrier”. (Participant 1)*

> *“I hope things will change now because we were provided with a motorbike, and we can go and collect the tests ourselves though it’s a long ride to the district.” (Participant 4)*

##### Power supply/internet technology

Other potential barriers, including pragmatic considerations such as availability of electricity and technological resources, remained persistent obstacles to implementation. Participants in most facilities expressed concern over lack of power supply, which affected the ability to maintain cold chain, and this affected the availability of POC diagnostic tests. For example, several participants mentioned that facilities had no electricity, and this limited the use of certain serum based syphilis rapid tests that require a centrifuge for blood separation.

> *“The challenge we face with POC testing and is a barrier is that there is no electricity, we don’t have enough solar power for a cold chain. Testing for syphilis and giving the mother results and treatment on the same day is not possible, because we must draw blood first and allow it to separate and this takes time, maybe the whole day, then we will do the test next day and by then the mother has gone. But if we had electricity and use a centrifuge which uses electricity, this problem would not be there. They should supply power and provide us with a centrifuge.” (Participant 11)*

> *“We don’t even have electricity we are using solar, this solar is not even enough to cater for the whole night … so I think this is a barrier.” (Participant 8)*

### Access to knowledge and information

Access to knowledge and information was highlighted as a facilitator that strongly encouraged implementation efforts. Facility providers at clinics reported having had training and sufficient knowledge on the rationale and use for POC testing interventions. They valued the initial training and orientation that was provided, which improved knowledge and promoted a supportive environment for implementation. Cited successful examples of training on POC testing included HIV cascade training through workshops, targeted mentoring of facilities and peer-to peer training, and this promoted willingness to adopt POC interventions. However, a few other informants stated low access to information such as printed manuals and materials such as protocols as a barrier to introducing more additional tests. They expressed a desire for regular training/capacity building to keep abreast with current HIV related knowledge and new technologies.

> *“We are happy we have had workshops and trainings on rapid tests for HIV, malaria, all rapid tests as part of management of these diseases and the training have been really good. It gave us an understanding of what was going to happen and what we were hoping to achieve.” (Participant 3)*

> *“It was quite a while ago, we have not had trainings or capacity building sessions for some time now and that is a challenge that we have.” (Participant 7)*

### Individual characteristics

Constructs examined under this domain were knowledge and belief, which were important facilitators. Healthcare providers at facilities were acquainted with knowledge and skills using POC testing to screen like HIV, malaria, BP measurements. The benefits of knowledge about the specific POC devices, the value and the rationale for using them seemed to highly motivate their work. This led informants to mention at least HIV, syphilis and malaria rapid tests as examples of POC tests with which they are familiar, and that more other tests should be offered at primary care level to improve health outcomes of patients and ultimately reduce referral and health expenditure in hospitals.

> *“I know HIV test kits, malaria test kits, syphilis and others for Hb and so on which are used in point of care. So in short my role is to mobilise commodities to make sure that they are made available and must be conducted daily on patients so that they receive suitable treatment and prevent diseases to escalate to higher levels and also reduce health care expenditure in hospitals” (Participant 9).*

Self-efficacy, referred to as the individual belief in their own capabilities to execute courses of action to achieve implementation goals, was found to have a positive presence as facilitator. Prior to the introduction of POC devices, facility providers lacked confidence in their individual abilities to conduct particular tests, e.g. HIV, but with orientation and guidance they expressed that they gained confidence and were excited to make a contribution to improving care in PHCs.

However, some personal attributes such as the user’s skills and competency were expressed by some as negative barriers. Regarding training, some participants expressed the view that the absence of regular training on POC testing made it harder to adopt the intervention, even when they had knowledge of other test devices available. Thus, the participants felt that on-going training programmes should be provided to increase their competencies to use diagnostic tests and could be a facilitator for implementation programmes.

> *“We need training in order for us to conduct more test for patients and this is going to help us a lot in our work but for now we have no guidance and this is a challenge.” (Participant 11)*

### Implementation process

Engagement as a construct within the process domain was found to positively affect implementation, particularly by those identified as champions. The involvement of nurses as champions drove the implementation forward due to their knowledge and professional role as front-line health providers. However, other facility-based health workers reported inadequacies in their involvement to support POC diagnostics integration. The reported insufficiencies included lack of decisions on selection and diagnostic test availability in primary healthcare facilities. The informants noted that they had no access to other tests, and they felt that they were not in an influential position to make decisions regarding test availability. Others, too, felt that they could not order the diagnostic tests beyond what a health post could handle.

> *It’s good that we as nurses, we were involved because we understand our role as professionals, and can make sure that we use the tests to care for our patients and treat them accordingly.” (Participant 3)*

> *“I think we are only required just to handle just a few of those tests, at this health level, am sure we can’t order something beyond a health post. We are not involved in decisions for the tests kits.” (Participant 2)*

## Discussion

The study sought to identify and understand factors that influence POC diagnostics implementation from the perspectives of implementing stakeholders in a positive and negative manner. The key drivers of implementation were examined systematically across the CFIR domains.

With regard to intervention characteristics, stakeholders reported numerous benefits of POC diagnostics interventions. Indeed, with regard to the relative advantage of timely return of results to improve patient management, POC testing created enthusiasm, uptake and desire for implementation of more and a wide range of POC diagnostic tests in PHCs to improve care. These findings support a growing body of evidence suggesting that POC testing can improve timely return of results and can save lives annually [9, 12, 24]. Some positive factors including leadership engagement, enthusiasm, knowledge and belief regarding the value of the intervention, self-efficacy, competence and confidence were drivers for successful implementation. In agreement with these results a recent study that evaluated at-birth POC testing in Kenya showed similar findings as enablers to implementation [24]. In addition, the process domain was also important. Key champions such as nurse practitioners in PHCs were critical for their role as frontline providers to move the implementation forward. Champions were found to facilitate PHC organisations’ and professionals’ decision to work with the intervention and were therefore important drivers of its success. The importance of champions has been described elsewhere to be effective in aiding implementation of evidence-based practice in healthcare settings [25].

While factors related to the intervention (perceived gains) and alignment to the national goals [26] are present, political commitment is a prerequisite for sufficient funding and a requirement for provision of commodities for sustainability and achieving desirable outcomes. The issue of limited financial resources is not such an unexpected finding. It is a potentially relevant sustainability factor to take into consideration when introducing any interventions in PHCs. Financial resources are important factors in any kind of process and they are frequently cited in models on the introduction of health interventions [27, 28]. Nevertheless, the importance of external funding partnerships was mentioned as critical enablers to implementation. However, both negative and positive effects of such partnerships have been reported elsewhere [29]. It is therefore important for national level implementers to be mindful when collaborating with stakeholders on global initiatives to ensure successful multi-sectoral collaboration.

Other important barriers in the outer setting were related to procurement and supply chain challenges leading to stock-outs. This finding aligns with a recent randomised trial in Zambia that examined the effect of three supply chain structures. It reflects the idea of supply chain system re-design, best management practices and routine system audit performance as crucial to reducing stock-outs of essential resources [30].

All barriers identified in the inner setting had a negative influence on implementation. Reporting of stock-outs is not surprising as this aspect is a common occurrence in LMICs [31], resulting in missed opportunities for testing. The qualitative results on workforce support the quantitative evidence from our parent study (unpublished) that suggests adequate health personnel to be essential for expediting POC testing in rural primary healthcare practices. Addressing workforce barriers is crucial to implementation support [29]. According to literature task-shifting, which is a challenge, may overcome the barrier of increased workload [30]. Other structural issues related to transportation, power supply, space limitations and technological connectivity will remain persistent threats to sustainable implementation. These findings further align with existing evidence from other studies on POC testing undertaken elsewhere, including sub-Saharan Africa [6, 9, 18, 31–35]. However, we recommend political determination and continued engagement among key stakeholders to alleviate some of these challenges.

The CFIR provided a useful structure to elicit key barriers and facilitators across domains, thus furthering our understanding of the implementation context of POC diagnostics services. The framework guided the results presentation, highlighting the CFIR domains and constructs that were most represented in the analysis. We did not specifically use the CFIR terminology to report our findings but rather framed our key findings in a language familiar to our readers. The CFIR is well established and used in a wide range of health-related implementation research, and its application is intended to be flexible [22, 36]. Thus, our reporting demonstrates its flexibility and versatility.

### Strengths and limitations

The study provides an in-depth examination of implementation factors, but due to the context-specific nature of the results, application in other settings should be done with caution. The study has several strengths, including inclusion of perspectives from several implementing stakeholders and use of a theoretical framework that guided the analysis to capture contextual factors influencing implementation in a rural context.

Despite these strengths, lack of direct patient input is a limitation. The study’s inclusion of implementing stakeholders and non-inclusion of patients might have swayed our results to over-represent the inner setting. Future studies should assess the perspectives of patients. Another limitation is that although the analysis was conducted through the lens of the CFIR, the interview guide was not designed to ask questions about specific CFIR constructs. However, the guide ensured that data collected was related to our research question.

### Conclusions

The study identified key contextual factors that supported and hindered the implementation of POC diagnostic interventions in PHCs. POC diagnostic innovations have proven potential for high impact intervention amidst implementation barriers and facilitators organised as per the CFIR framework. Indeed, addressing barriers at multiple levels is critical for successful implementation. The drivers of implementation identified can inform future efforts, sustainability and scale-up of POC interventions in rural settings.

## Data Availability

All data produced in the present work are contained in the manuscript

## Acknowledgements

The authors would like to thank the Ministry of Health of Zambia at the national, provincial, and district levels, as well as the private health sector, for their relevant roles to support this study. The authors would like to also thank all stakeholders who participated in the study. The authors sincerely acknowledge SANTHE for the generous financial support, the College of Health Sciences, University of KwaZulu-Natal, Discipline of Public Health, for providing the space and resources, and the University of Zambia for rendering their support to the study.

